# Early Palliative Care Intervention Helps with Decreased Length Of Stay And Cost Cutting In A Safety Net Hospital In The Central Valley Of California

**DOI:** 10.1101/2020.08.05.20169235

**Authors:** Soujanya Sodavarapu, Siamak M. Seraj, Gurinder Ghotra, Malkinder Singh, Nasim Khosravi, Kinnari Parikh, Syung Min Jung

**Affiliations:** Department of Internal Medicine, San Joaquin General Hospital, French Camp, CA-95231 Email; Department of Internal Medicine, San Joaquin General Hospital, French Camp, Ca-95231; Department of Internal Medicine, Department of Palliative Care, San Joaquin General Hospital, French Camp, Ca-95231

## Abstract

**Objective:** To determine if early palliative care intervention within two days of hospital admission affects the length of stay and cost savings.

**Methods:** Using a retrospective chart review, 570 patients who received palliative care consultation were reviewed between 2016 and 2018. 287 patients were seen within two days of days, and the total 355 were seen within three days of admission. Data on length of stay and total charges were analyzed for both groups.

**Results:** In the early consult group, both lengths of stay and cost of care in total charges decreased by 64% (p<0.0001) and 58% (p<0.0001), respectively. Multiple linear regressions showed everyone day increase in the date of the consultation is associated with an increase in the length of stay by 1.02 days. (R-squared 0.65, p-value <0.0001, CI 0.95-1.09). The number of palliative care consultations increased by 60% from 2016 to 2017.

**Conclusion:** Our study reiterates the importance of a multidisciplinary approach in identifying patients who will benefit from palliative care consultation and addressing goals of care early in their hospital course. As such, our study suggests the importance of emphasizing early palliative care and its potential benefits in public hospitals.

## Introduction

There have been a lot of technological advancements in medicine, but they do not necessarily always translate to quality of care and psychosocial wellbeing of the patients and their families [1]. Palliative care has emerged as a specialty that addresses quality of life along with the emotional, psychosocial and physical wellbeing of critically ill patients, but also helps the burden of the health care system [1,2].

The implementation of patient-centered palliative care teams in the US has increased to 178% from 2000 to 2016 in hospitals with more than 50 beds [3]. This has been attributed to the improved quality of care implements seen with palliative care.

With approximately 90 million Americans living with severe illness and multi-morbidities, this number is expected to more than double over the next 25 years [2]. Medicare spending increased to $651 billion in 2019 from $438 billion in 2013. 17% of patients with six or more comorbidities account for 53% of annual spending [4]. 25% of Medicare's annual spending is used by patients during the last 12 months of their lives.

The following is a study of the multidisciplinary palliative care team approach implemented at the only safety-net hospital in the central valley of California, which measures the impact of the early palliative intervention on the cost of care and length of stay.

## Methods

Ours is a central valley safety net hospital in which a dedicated multidisciplinary palliative care team was formed in 2015. It consisted of board-certified palliative care specialists, palliative care clinical nurses, social workers, and discharge planners. Discharge planners have been trained to screen patients for potential palliative care consultation and determine eligibility based on admission diagnosis. Screening criteria for the discharge planners were patients with advanced cancer with documented Stage 3 or 4 cancer, having advanced organ failure (Congestive heart failure (CHF) with ejective fraction (EF) < 30%, Chronic obstructive pulmonary disease (COPD) on home oxygen, End-stage renal disease (ESLD)) and patients with advanced dementia. Primary teams have since been encouraged early palliative care consultation if the criteria are met.

The objective of our study was to determine if early palliative care consultation had an impact on the hospital charges and its cost-effectiveness for early vs. later intervention. Determine the change in the length of stay, and to determine the percentage of patients discharged home with palliative care/hospice with early vs. later intervention.

We conducted a retrospective chart review from January 2016 through June 2018, who received palliative care consultation during their admission to our hospital. We included patients age 18 years and older. We analyzed patients' age, sex, primary and secondary diagnosis at the time of admission, comorbidities, insurance, length of the stay, total changes of the hospital stay, discharge with or without hospice, and patients who transferred on comfort care. We analyzed patients within two days and three days of admission with patients who received consultation after 2 and 3 days of admission.

Hospital costs were obtained from the hospital finance department and included the charges for medications, procedures, the services provided to the patient, and the billing by the physicians who cared for the patients.

## Results

We reviewed 562 patient charts from January 2017-June 2018, who received palliative consultation during the hospital admission. 287 patients were seen within two days of days, and the total 355 were seen within three days of admission. We further analyzed the patients in each group having a Palliative Performance Scale (PPS) score of <50 and >50. A PPS score of >50 is considered a good prognostic indicator, and a PPS score <50 is considered a poor prognostic indicator. 200 (47.69%) patients had a PPS score of >50, and 363(64.41%) of patients had a PPS score <50. We also found that the total number of consults in our hospital was 165 in 2016 and 265 in 2017, which is a 60% increase, which showed a good utilization of the multidisciplinary team which formed in 2015.

Of the 562 patients, 168(29.89%) were females, and 394(70.11%) were males. Further subdivision of the diagnosis showed that most of the patients either had cancer or cardiovascular diagnosis, which consisted of 53% of the patients (chart 1). The mean age of presentation was 64.2 ± 14.0 years.

**Chart 1:**
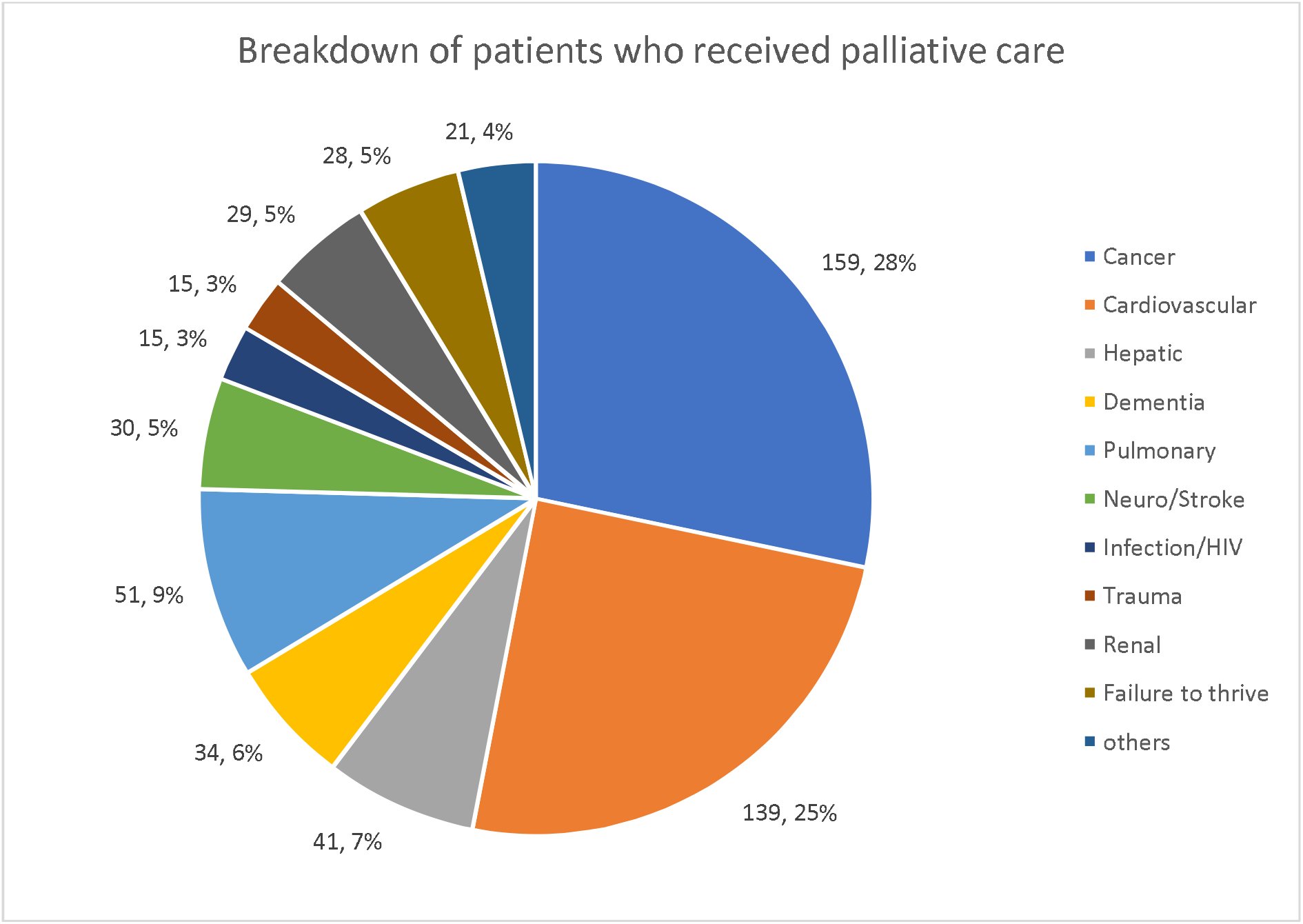
Breakdown of diagnosis of patients receiving palliative care.

Insurance in our patient population was as follows: Medicare/Medicare-Medical (n=180) 31.6%, Medical/HPSJ (n=208) 36.5% and Others (n=182) 31.9%. Of all the patients given consult in our hospital in this study, 47.4%(n=270) patients were discharged with hospice, 38.6% (n=220) were discharged without hospice and 80 (14%) patients were initiated on comfort care.

Length of stay of patients at two days consultation was 3.3 days vs. 9.1 days for patients who received consultation after two days with statistical significance. Similarly, the length of stay of patients at three days consultation was 3.5 days vs. 10.7 days after three days of consultation and was statistically significant (Table 1) Analyzing the total charges for patients palliative care at two days showed a mean of $77,298.5 vs. $185,912 for patients who received palliative care consult after two days was statistically significant. Similarly, total charges at three days vs. after three days consultation showed total changes mean of $80,817.5 vs. $218,088 for patients receiving care after three days of hospitalization is statically significant (Table 2).

**Table 1:**
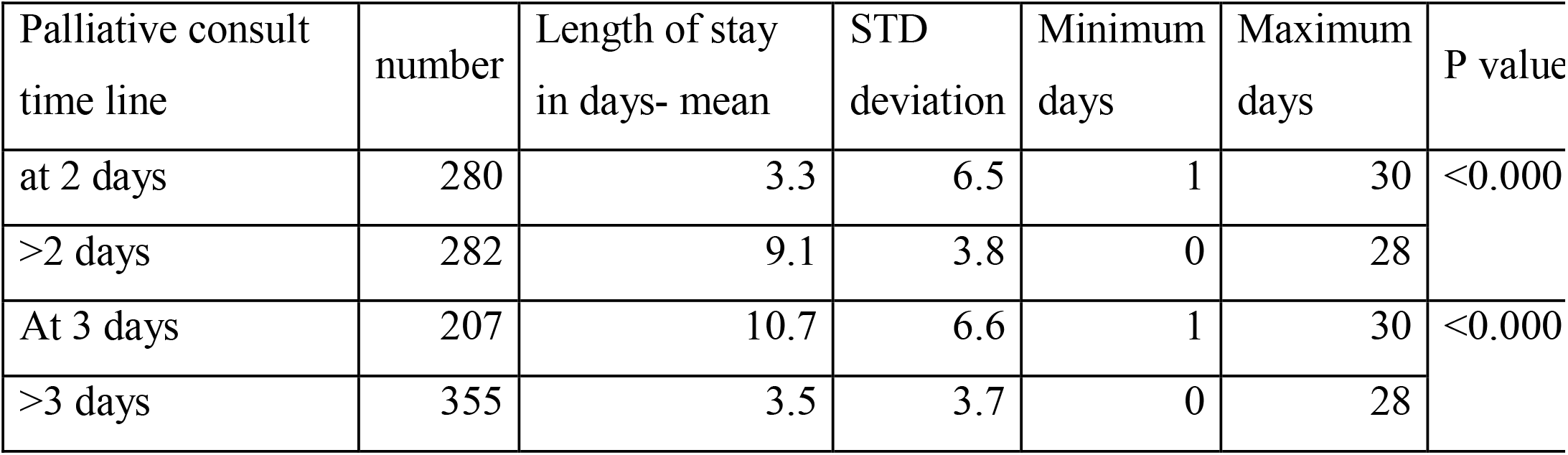
Length of stay for consultation at 2 and 3 days of admission.

**Table 2:**
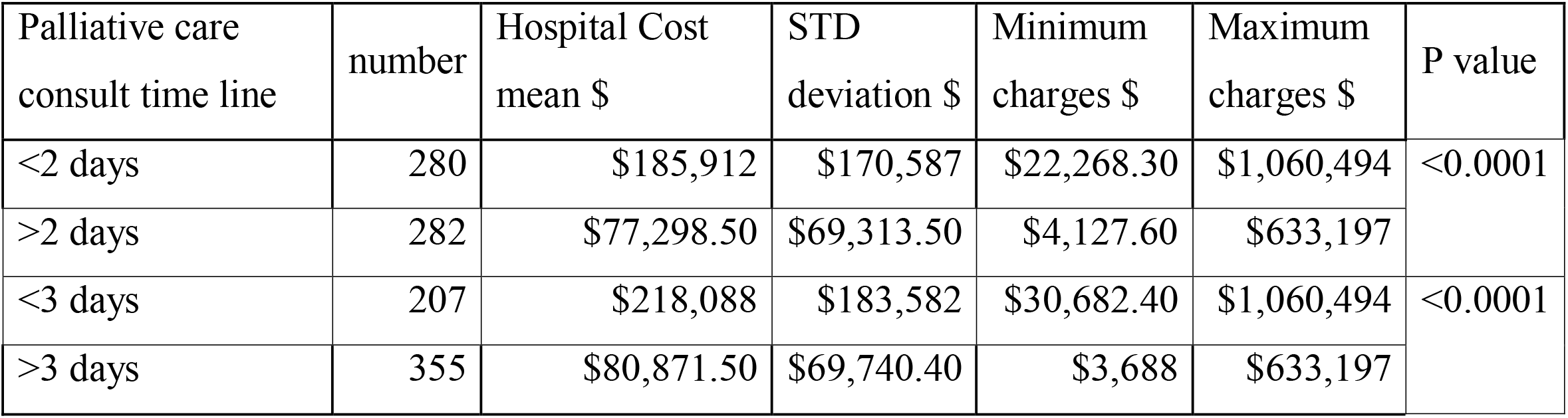
Hospital charges at 2 days and 3 days of admission.

| Palliative care consult time line | number | Hospital Cost mean \$ | STD deviation \$ | Minimum charges \$ | Maximum charges \$ | P value |
| --- | --- | --- | --- | --- | --- | --- |
| <2 days | 280 | \$185,912 | \$170,587 | \$22,268.30 | \$1,060,494 | <0.0001 |
| >2 days | 282 | \$77,298.50 | \$69,313.50 | \$4,127.60 | \$633,197 | |
| <3 days | 207 | \$218,088 | \$183,582 | \$30,682.40 | \$1,060,494 | <0.0001 |
| >3 days | 355 | \$80,871.50 | \$69,740.40 | \$3,688 | \$633,197 | |

We compared the number of patients with PPS scores <50 vs.>50 for patients who received palliative consult and found that 200 (35.59%) of patients had PPS score >50 and 363 (64.41%) patients had PPS score of <50. Of all the patients who received hospice (n=294), 39 has a PPS score of >50, and 255 patients had a PPS score of <50, which was statistically significant with a p-value of <0.001.

Analyzing the PPS score in patients before two days and after two days of the consultation showed the following results (Table 3).

**Table 3:** PPS score in patients before 2 days and after 2 days of the consultation

| Consult timeline | PPS score | total patients | percentage of patients | P value |
| --- | --- | --- | --- | --- |
| <2 days | >50 | 111 | 19.75% | 0.0607 |
| <2 days | <50 | 171 | 30.43% |  |
| >2 days | >50 | 89 | 15.84% |  |
| >2 days | <50 | 191 | 33.99% |  |

Multiple linear regressions showed every 1-day increase in the date of admission to date of the consultation is associated with an increase in the length of stay by 1.02 days. (R-squared 0.65, p-value <0.0001, CI 0.95-1.09).

## Discussion

The increasing costs of health care not only affects the individuals and families but the overall economic health of the country [5]. According to Wammes, the sickest and the most complicated patients use 30% of health care costs in the last year of their life span [6]. Many studies have shown that patients with advance life-threatening illness suffer from untreated pain, have lengthy hospital stays, often have costly medical treatments which are usually low yield and unlikely to change the outcome of the course and overall provides lower satisfaction to the patient and families around the end of patient's life [7].

As physicians or caregivers treating the terminally ill patient when no cure was possible, and end of life was inevitable, the aim is to reduce patient suffering. The field of palliative care medicine grew out of the hospice movement in the 1960s [8]. The core principles of palliative care approach are to improve the quality of life for both patients and their families facing the end of life terminal illness. Palliative care provides relief from pain and distressing symptoms, affirms life and regards dying as a normal process, intends neither hasten nor postpone death, integrates the psychological and spiritual aspects of patient care, and offers a support system to help patients live as actively as possible until death [9].

Palliative care program involves interdisciplinary teams consisting of trained physicians, nurses, social workers, and discharge planners who collectively can help form a bridge between improving the quality of care of the sickest patients and as well decrease the overall cost of treatment for the terminally ill. [5]

There are many ways the palliative care approach leads to cost savings in terminally ill hospitalized patients. For example, a reduced cost of overall hospitalization can occur from reduced length of stay (LOS) or from a less intense treatment approach in terminally ill or a combination of both [10].

Our results demonstrated that the direct cost of the hospitals could be significantly reduced for patients with significant comorbidities when timely palliative care consultation is obtained. As a result, you have reduced utilization of the facility, reduced number of unnecessary procedures which are unlikely to change the overall outcome, and hence reduced length of stay in the hospital. Literature has shown that early inpatient palliative care consultation reduced the length of hospital stay by about 30% and the cost of care by 22% to 33% [11-14].

Most of the research done to date has compared palliative care consultation to usual care. A recent prospective trial done by Mary P et al., published in 2017, showed that there were 63% cost savings in patients who had palliative care intervention compared to the usual care [15]. A metanalysis of 6 research studies, including both retrospective and prospective studies, have demonstrated that palliative care consultation resulted in cost savings from $2,666 to $9,237 when compared to the usual care group [16].

A study published in 2014 compared palliative consultation within ten days of admission versus consultation obtained greater than ten days of admission found to have significant cost savings in patients who died in the hospital compared to patients who were discharged [17]. Another prospective study compared palliative care consultation within six days of admission and two days of admission for cancer patients and found 14% cost reduction if a consultation was obtained within six days and 24% cost reduction if a consultation was obtained in 2 days compared with no intervention.

Most of these studies discussed above have shown the reduction in hospital costs but did not comment much on the length of stay of patients who had a palliative intervention. However, there have been initial studies which have shown positive economic impact but found no association between palliative care consult team (PCCT) and length of stay (LOS), leading to the inference that all cost-savings have been through the reduced intensity of care: Patients who received palliative care stayed as long in hospital as matched cohorts but utilized lesser services during their admission reducing the cost [18].

A multicenter prospective study published by May et al. in 2015 on palliative care intervention in advanced cancer patients showed that palliative care intervention was effective in reducing the length of stay when the intervention happed at two days of admission rather than consultation happening at days 6, 10 or 20 of admission; which is among the very few studies published that showed the significance of the decreased length of stay to hospital costs [19].

One study similar to our study that was done in New York, published in 2018, compared early palliative care consultation within three days of admission versus later palliative care interventions greater than three days of admission and found to have an average length of stay of 6.09 vs. 16.5 days, respectively [18]. Of note, the study compared and looked at the Palliative Performance Scale (PPS) of < 30, 30-60, and >60.

Our study is unique as it compared the studies currently available in the literature on the early palliative care approach in several aspects. Most of the studies have compared palliative care intervention to usual care. Our study compared various aspects of palliative intervention in patients with early palliative care consultation within two days of admission versus after two days of admission. Our study is a retrospective study from January 2016 to June 2018, which looked at all patients (562 patients) who received palliative care consultation during their hospitalization. Of the 562 patients, 282 patients were seen by the palliative care consultation team (PCCT) during the first two days of their hospitalization and 355 patients at three days of admission.

Our results demonstrated that sooner the palliative care consultation is obtained in a terminally ill patient with multiple comorbidities, the reduced length of stay, and reduced hospital cost. For example, the average length of stay for patients receiving PCCT within two days was 3.3 (p-value< 0.0001), whereas 9.1 LOS for PCCT greater two days. However, patients receiving PCCT on day three still had a LOS of 3.5 (p-value <0.0001) compared to patients receiving consultation greater than three days. Similarly, the cost for patients receiving consultation within 2 days was $ 77, 298 (p-value < 0.0001) versus $ 185, 912 (p-value <0.0001) for patients obtaining PCCT greater than 2 days. However, patients who received PCCT at day 3 had cost of $ 80, 871 (p-value< 0.0001). In conclusion, our data showed that PCCT intervention was done within two days of admission resulted in reduced length of stay as well as reduced hospital cost in terminally ill advanced disease patients admitted to the hospital. However, if the PCCT was obtained on day 3, it was still better in achieving a lesser length of stay and cost savings compared with PCCT done greater than three days. Hence, sooner the PCCT can be obtained, the reduced length of stay and hospital cost in our palliative care patient population.

We further analyzed the patients in each group having a Palliative Performance Scale (PPS) score of <50 and >50. A PPS score of >50 is considered a good prognostic indicator, and the PPS score <50 is considered a poor prognostic indicator. Of our patients, 200 (47.69%) patients had PPS score of >50 and 363 (64.41%) patients had PPS score <50.

## Conclusion

Palliative care consultation within two days showed a decreased length of stay in hospital and hence lesser total charges when compared to patients who had consultation after two days, and the difference is statistically significant. In addition, patients with PPS scores >50 were higher in the group with palliative care consultation within two days. Our study reiterates the importance of a multidisciplinary approach in identifying patients who will benefit from palliative care consultation and addressing goals of care early in their hospital course. As such, our study suggests the importance of emphasizing early palliative care and its potential benefits in public hospitals.

## Data Availability

The data that support the findings of this study are available on request from the corresponding author. SS

## Contribution

SS, GG contributed to the drafting the manuscript; MS, NK, helped in data collection, SMS did the statistical analysis for the project, KP, SMJ critically revised the manuscript for intellectual content, SMJ had given the intellectual input and supervised the work, and all authors approved the final version of the manuscript for publication.

## References

1. Davis MP, Temel JS, Balboni T, Glare P. A review of the trials which examine early integration of outpatient and home palliative care for patients with serious illnesses. Ann Palliat Med. 2015;4(3):99D 121. doi:10.3978/j.issn.2224-5820.2015.04.04

2. Welcome to the California Hospice and Palliative Care Association. (2020, August 1). Retrieved from https://www.calhospice.org/

3. Palliative Care Continues Its Annual Growth Trend, According to Latest Center to Advance Palliative Care Analysis. (2020, August 1). Retrieved from https://www.capc.org/about/press-media/press-releases/2018-2-28/palliative-care-continues-its-annual-growth-trend-according-latest-center-advance-palliative-care-analysis/

4. Multiple Chronic Conditions. (2020, August 1). Retrieved from https://www.cms.gov/Research-Statistics-Data-and-Systems/Statistics-Trends-and-Reports/Chronic-Conditions/MCC_Main

5. Morrison RS, Dietrich J, Ladwig S, Quill T, Sacco J, Tangeman J, Meier DE. Palliative Care Consultation Teams Cut Hospital Costs For Medicaid Beneficiaries. Health Affairs. 2011; 30(3), 454-463. doi: 10.1377/hlthaff.2010.0929

6. Wammes, JJG, van der Wees PJ, Tanke MAC, Westert GP, Jeurissen PPT. Systematic review of high-cost patients’ characteristics and healthcare utilisation. BMJ Open. 2018; 8(9), e023113. doi:10.1136/bmjopen-2018-023113.

7. Morrison RS. (2008). Cost Savings Associated With US Hospital Palliative Care Consultation Programs. Archives of Internal Medicine. 2008;168(16), 1783. doi:10.1001/archinte.168.16.1783

8. Clark, D. From margins to centre: a review of the history of palliative care in cancer. The Lancet Oncology. 2007; 8(5), 430–438. doi:10.1016/s1470-2045(07)70138-9

9. WHO Definition of Palliative Care. (2020, August 1). Retrieved from http://www.who.int/cancer/palliative/definition/en.

10. Fitzpatrick J, Mavissakalian M, Luciani T, Xu Y, Mazurek A. Economic Impact of Early Inpatient Palliative Care Intervention in a Community Hospital Setting. Journal of Palliative Medicine. 2018; 21(7), 933–939. doi:10.1089/jpm.2017.0416

11. Norton SA, Hogan LA, Holloway RG, Temkin-Greener H, Buckley MJ, Quill TE. Proactive palliative care in the medical intensive care unit: effects on length of stay for selected high-risk patients. Crit Care Med. 2007;35(6):1530–1535. doi:10.1097/01.CCM.0000266533.06543.0C

12. May P, Garrido MM, Cassel JB, et al. Prospective Cohort Study of Hospital Palliative Care Teams for Inpatients With Advanced Cancer: Earlier Consultation Is Associated With Larger Cost-Saving Effect. J Clin Oncol. 2015;33(25):2745–2752. doi:10.1200/JCO.2014.60.2334

13. Ciemins EL, Blum L, Nunley M, Lasher A, Newman JM. The Economic and Clinical Impact of an Inpatient Palliative Care Consultation Service: A Multifaceted Approach. Journal of Palliative Medicine. 2007;10(6), 1347—1355. doi:10.1089/jpm.2007.0065

14. Cassel JB, Kerr K, Pantilat S, Smith TJ. Palliative care consultation and hospital length of stay. J Palliat Med. 2010;13(6):761–767. doi:10.1089/jpm.2009.0379

15. May P, Garrido MM, Cassel JB, et al. Cost analysis of a prospective multi-site cohort study of palliative care consultation teams for adults with advanced cancer: Where do cost-savings come from?. Palliat Med. 2017;31(4):378–386. doi:10.1177/0269216317690098

16. May P, Normand C, Cassel JB, et al. Economics of Palliative Care for Hospitalized Adults With Serious Illness: A Meta-analysis. JAMA Intern Med. 2018;178(6):820–829. doi:10.1001/jamainternmed.2018.0750

17. McCarthy IM, Robinson C, Huq S, Philastre M, Fine RL. Cost Savings from Palliative Care Teams and Guidance for a Financially Viable Palliative Care Program. Health Services Research. 2014;50(1), 217–236. doi:10.1111/1475-6773.12203

18. May P, Garrido MM, Cassel JB, et al. Prospective Cohort Study of Hospital Palliative Care Teams for Inpatients With Advanced Cancer: Earlier Consultation Is Associated With Larger Cost-Saving Effect. J Clin Oncol. 2015;33(25):2745–2752. doi:10.1200/JCO.2014.60.2334

19. Luckett T, Phillips J, Agar M, Virdun C, Green A, Davidson PM. Elements of effective palliative care models: a rapid review. BMC Health Serv Res. 2014;14:136. Published 2014 Mar 26. doi:10.1186/1472-6963-14-13

